# The descriptive epidemiology of type 2 diabetes in the United Kingdom from 2004 to 2021

**DOI:** 10.1101/2024.03.04.24303693

**Authors:** Craig J. Currie, Jessica G. Currie, Sarah E. Holden, Christopher Ll. Morgan, Benjamin R. Heywood, John R. Peters, the Livingstone Development Team

## Abstract

**Purpose:** To provide contemporary estimates of the incidence and prevalence of type 2 diabetes in the UK.

**Methods:** Data from UK primary care (CPRD) were analyzed using an automated, analytical platform that produces validated, rapid analytics–Livingstone.

**Results:** We selected 1,125,028 people with type 2 diabetes (44.7% female). The crude incidence was stable. In 2004 the incidence rate was 4.18 cases per 1,000 people, reducing to 4.13/1,000 in 2021. There was a shift in age-specific incidence to earlier age-groups. The crude prevalence showed a marked increase, rising from 2.95% to 5.41%, levelling off slightly in later years. Increases in prevalence were observed in all groups. Northern Ireland consistently had the lowest prevalence, and Wales had a notably higher prevalence, increasing from 3.71% in 2004 to 6.37%. The mean age at diagnosis decreased in females by 5.5 years to 58.5 years, and by 2.5 years in men, to 58.8 years.

**Conclusions:** The incidence of type 2 diabetes was largely stable over 20 years but there were changes over time in age-specific groups resulting from a shift to earlier onset or earlier diagnosis. The shift to earlier diagnosis was far more pronounced in women than in men. Prevalence increased markedly, but the rate of increase appeared to be leveling-off a little. There were notable differences in the epidemiology of type 2 diabetes between the four constituent countries of the UK. The number of younger people diagnosed with type 2 diabetes remains a concern.

**Guarantor:** Craig Currie is the guarantor of this work and, as such, had full access to all the data in the study and takes responsibility for the integrity of the data and the accuracy of the data analysis.

## Introduction

Increases in recorded prevalence of a condition may be due to both a genuine increase in incidence; artefact (for example increased surveillance or screening), changes in demographics, or changes in case definition. As life expectancy increases, a larger proportion of patients live long enough to develop age-related conditions and also existing cases live longer.^1^

The pattern of the various risk factors predisposing to the onset of type 2 diabetes in the general population shifts continually. Regarding type 2 diabetes, the two main risk factors are the age structure of the population and the pattern of body composition. The risk of type 2 diabetes increases with increasing age, and with increasing obesity. Other factors contribute to incident risk, for instance: sex, lifestyle, and ethnicity.

In the United Kingdom (UK) there is an increasing absolute frequency and proportion of older people in the population. For instance, the number of people aged 85 and over, which was 1.7 million in 2020 (2.5% of the population), is expected to nearly double to 3.1 million by 2045, representing 4.3% of the UK population^2^. On the other hand, by mid-2030, the number of children (0 to 15 years) is projected to decrease by 1.1 million.

Over the past two decades, the level of obesity in the UK has seen a worrying escalation. By 2030, the UK is expected to see a further rise in obesity levels. Projections suggest 11 million more obese adults, potentially leading to an increase in associated health problems^3^. These worsening patterns of risk inevitably have resulted in a corresponding increase in the burden of type 2 diabetes and its related sequalae.

The objective of this study was to characterize the descriptive epidemiology of type 2 diabetes in the UK from 2004 until 2021. Continuous updates of these incidence and prevalence metrics are necessary to maintain an understanding of the evolving impact of this increasingly common disease. Unusually, we also present differing estimates for the four nations that comprise the UK. Using the same data source, but using more routine epidemiological methods of data analysis, our most recent estimate for type 2 diabetes prevalence in the UK was published in 2017, characterizing the prevalence rate in years 1991 to 2013^4^. We reported that the crude population prevalence in 1991 was only 1.4%, increasing to 4.5% in 2013. Again, using the same data source, our most recent estimate of the incidence of type 2 diabetes was published a little earlier, in 2013, and related to incidence rates in 1991 to 2010^5^. In 2010, the crude incidence rate we reported was 0.52%, increasing from 0.17% in 1991.

## Methods

Livingstone is an automated analytical platform that produces rapid analysis of linked, UK National Health Service (NHS) data, including a validated, descriptive epidemiology module. Detailed methods that underpin the descriptive epidemiological statistics produced by Livingstone, and validation of these estimates have been published recently^6^. The following description is a summary.

### Data source

The source of NHS data was the Clinical Practice Research Datalink (CPRD). CPRD evolved from the General Practice Research Database (GPRD) and harnessed anonymized patient level data. It offers researchers access to comprehensive primary and linked secondary healthcare data, including information on prescribed drugs and laboratory data. For this study we utilized only data from general practice, from GOLD (Vision) and Aurum (EMIS). Patient migration between practices using these two systems was accounted for by excluding migrated practices from GOLD.

### Case identification

People were classified as having a diagnosis of type 2 diabetes if they had a relevant clinical diagnosis recorded anywhere in their case history. A second set of search criteria selected cases where there was a non-specific diagnostic code for diabetes, but the person had received a prescription of an anti-hyperglycemic drug other than insulin within two years of recorded diagnosis. Checks were made to assure that women with gestational diabetes were excluded. A full list of the diagnostic codes used here is provided at Supplementary Table 1. Whilst it is possible that patients with type 2 diabetes can revert to a state of diabetes remission, this is rare and usually transient, therefore a diagnosis was treated as a lifelong event.

### Annual incidence

Inclusion for the incidence analysis required patients in the diabetes cohort to have had an incident diagnosis date within the CPRD follow-up period within a specific year. This initial diagnosis had to be at least 90 days after their GP registration date. The denominator for the incidence analysis included all patients of acceptable research quality in the dataset over the specific period being reported, with a minimum of 90 days’ registration (person time).

### Point prevalence

The point prevalence was determined. The base population for this prevalence study included all patients of acceptable research quality as defined by CPRD who had CPRD follow-up coinciding with the mid-year point. This included people who were registered, but they had not necessarily had a record of an appointment with their GP on the specific day of prevalence being reported. A type 2 diabetes case would be considered prevalent if they had a relevant diagnosis recorded in their clinical history prior to (inclusive) of the day of prevalence estimation.

### UK nationality

Region of residence is recorded in CPRD, including whether a GP practice was located in England (83.9% of cases), Northern Ireland (1.9%), Scotland (8.0%), or Wales (6.1%). Where necessary, the denominators were adjusted to account for geographical location.

### Statistical methods

Age and sex-specific rates were calculated, and the denominators used were appropriate to the numerator. Otherwise, no advanced statistical methods were used in this study that were not already detailed in the Livingstone epidemiology validation study^6^.

### Study approval

CPRD RDG approval was granted for this study (22_001774).

## Results

### Type 2 diabetes cases

We selected 1,125,028 people with a record of a clinical diagnosis of type 2 diabetes. Of these cases, 502,341 (44.7%) were female. Their mean body mass index (BMI) at diagnosis was 32.4 kg/m^2^ with an average blood pressure (BP) at diagnosis of 137/80 mmHg; mean total cholesterol, 5.1 mmol/L; mean high density lipoprotein (HDL), 1.2 mmol/L; mean low density lipoprotein (LDL), 3.0 mmol/L; mean triglyceride 2.3 mmol/L; and their mean estimated glomerular filtration rate (eGFR), 78.8 ml/min/1.73m^2^. At diagnosis, 19.7% were recorded as being smokers, and 42.6% ex-smokers. These people were often treated for some of these clinical issues.

### Age and sex distribution at diagnosis

The mean age at diagnosis over the whole period was 60.3 years old. However, the pattern at diagnosis showed marked variation by age and by sex (Figure 1). The mean age at diagnosis in 2004 was 62.5 years, reducing to 58.6 years in 2021; an overall average change of 3.9 years. These changes differed between males and females. The corresponding values in females were 64.0 and 58.5 years (Δ=5.5 years), and males, 61.3 years and 58.8 (Δ=2.5 years; Figure 1).

**Figure 1.**
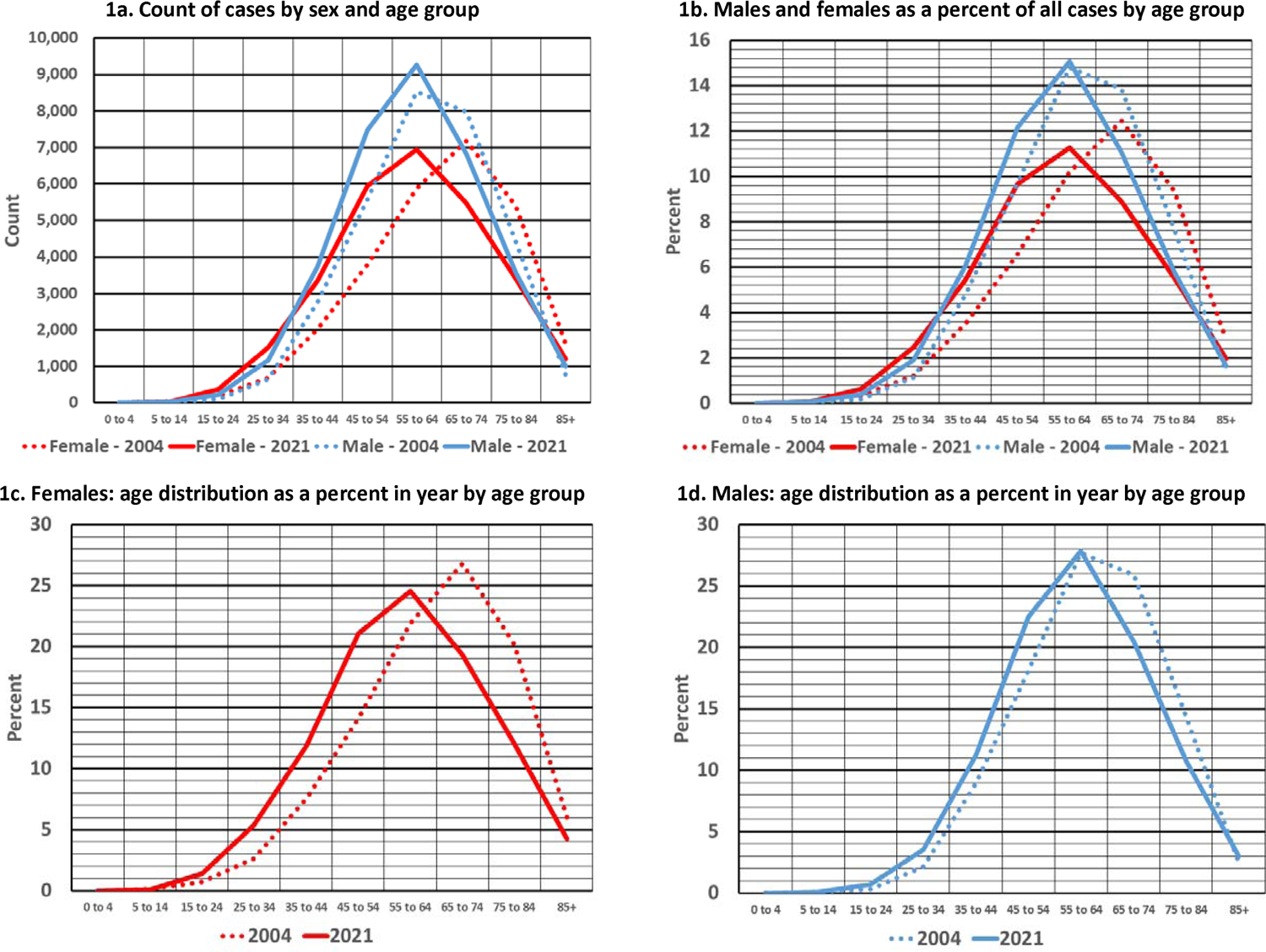
Age and sex distribution of people diagnosed with type 2 diabetes.

### Incidence

The overall, crude incidence rate over time showed little variation over the period (Figure 2a), except at the time of the COVID19 pandemic. In 2004 the crude incidence rate was 4.18 cases per 1,000 people, and this had reduced slightly to 4.13/1,000 people in 2021. There was a drop in 2020 to 3.11/1,000 people, that was likely to be COVID19 related. The highest recorded crude incidence rate was in 2004.

**Figure 2.**
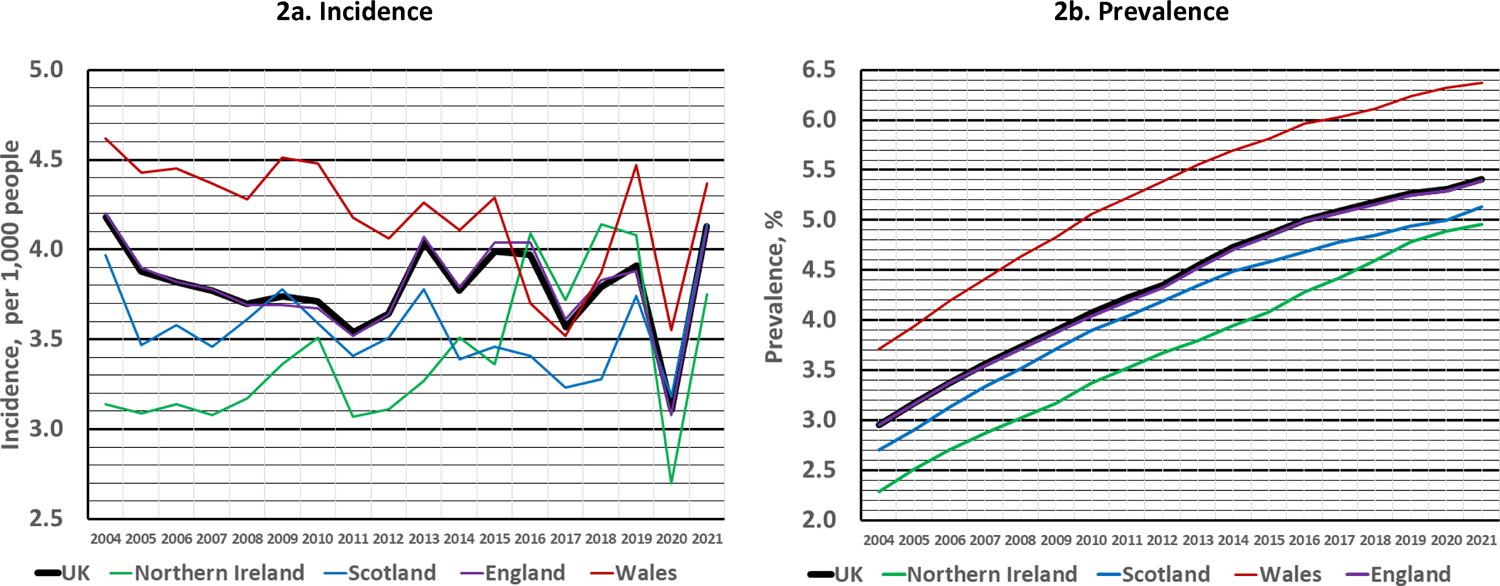
Crude incidence and crude prevalence of type 2 diabetes, by calendar year and by UK nation.

There were notable differences in the incidence of type 2 diabetes by UK nation (Figure 2a). In general, Northern Ireland had a lower incidence rate, and Wales had a higher incidence rate in earlier years, but the incidence rates tended to converge in later years. There was a marked drop in the incidence rate of all four nations in 2020.

As an example of the differential incidence rate in males over females, in 2021 men had a crude type 2 diabetes incidence rate of 4.47/1,000 men, whereas females had an incidence rate of 3.79/1,000 women (Table 1). The incidence rate ratio of males to females changed little over time from 1.14 in 2004 to 1.18 in 2021.

**Table 1.** Incidence rate per 1,000 people by year, sex, and age groups.

| Group | 2004 | 2005 | 2006 | 2007 | 2008 | 2009 | 2010 | 2011 | 2012 | 2013 | 2014 | 2015 | 2016 | 2017 | 2018 | 2019 | 2020 | 2021 |
| --- | --- | --- | --- | --- | --- | --- | --- | --- | --- | --- | --- | --- | --- | --- | --- | --- | --- | --- |
| All UK | 4.18 | 3.88 | 3.82 | 3.77 | 3.70 | 3.74 | 3.71 | 3.54 | 3.64 | 4.04 | 3.77 | 3.99 | 3.97 | 3.57 | 3.79 | 3.91 | 3.11 | 4.13 |
| Male | 4.46 | 4.18 | 4.16 | 4.15 | 4.11 | 4.23 | 4.22 | 3.99 | 4.10 | 4.48 | 4.15 | 4.39 | 4.39 | 4.01 | 4.22 | 4.33 | 3.40 | 4.47 |
| Female | 3.90 | 3.57 | 3.48 | 3.39 | 3.29 | 3.24 | 3.19 | 3.09 | 3.18 | 3.60 | 3.38 | 3.58 | 3.54 | 3.13 | 3.37 | 3.49 | 2.81 | 3.79 |
| 0-4 | 0.00 | 0.00 | 0.00 | 0.00 | 0.00 | 0.00 | 0.00 | 0.01 | 0.00 | 0.00 | 0.00 | 0.00 | 0.00 | 0.00 | 0.00 | 0.00 | 0.00 | 0.00 |
| 5-14 | 0.04 | 0.03 | 0.04 | 0.03 | 0.03 | 0.03 | 0.02 | 0.02 | 0.02 | 0.02 | 0.03 | 0.02 | 0.03 | 0.03 | 0.03 | 0.03 | 0.03 | 0.04 |
| 15-24 | 0.19 | 0.20 | 0.19 | 0.20 | 0.19 | 0.20 | 0.20 | 0.21 | 0.22 | 0.26 | 0.28 | 0.31 | 0.31 | 0.29 | 0.28 | 0.30 | 0.27 | 0.35 |
| 25-34 | 0.74 | 0.75 | 0.73 | 0.75 | 0.80 | 0.81 | 0.83 | 0.81 | 0.88 | 0.97 | 1.01 | 0.99 | 1.06 | 1.01 | 1.03 | 1.05 | 1.01 | 1.27 |
| 35-44 | 2.19 | 2.11 | 2.20 | 2.22 | 2.37 | 2.41 | 2.51 | 2.49 | 2.64 | 2.86 | 2.79 | 2.96 | 3.01 | 2.86 | 2.89 | 3.02 | 2.54 | 3.41 |
| 45-54 | 5.19 | 5.04 | 4.99 | 5.12 | 5.15 | 5.21 | 5.23 | 5.11 | 5.23 | 5.61 | 5.44 | 5.80 | 5.89 | 5.49 | 5.85 | 6.01 | 5.02 | 6.77 |
| 55-64 | 8.87 | 8.14 | 8.05 | 8.14 | 8.04 | 8.20 | 8.17 | 7.83 | 7.97 | 8.68 | 8.00 | 8.46 | 8.45 | 7.59 | 8.04 | 8.3 | 6.89 | 8.84 |
| 65-74 | 12.80 | 11.73 | 11.38 | 10.92 | 10.29 | 10.22 | 9.99 | 9.30 | 9.32 | 10.30 | 9.09 | 9.50 | 9.15 | 8.15 | 8.70 | 8.96 | 6.59 | 9.16 |
| 75-84 | 11.76 | 10.57 | 10.39 | 9.88 | 9.28 | 9.49 | 9.00 | 8.29 | 8.78 | 10.43 | 9.18 | 9.88 | 9.56 | 7.99 | 8.78 | 8.87 | 6.14 | 8.01 |
| 85+ | 7.92 | 7.14 | 6.82 | 6.18 | 5.98 | 5.72 | 5.67 | 5.32 | 5.54 | 6.85 | 6.90 | 7.19 | 7.32 | 6.13 | 6.81 | 7.20 | 4.89 | 6.39 |

In 2004, the highest incidence rate was observed in people aged between 65 to 74 years old (12.80 cases per 1,000 people, Table 1). However, the incidence rate in this age-group declined to 9.16/1,000 in 2021. It was 8.96/1,000 in 2019, pre-COVID19. There was an increase in the incidence rate between 2004 and 2021 in age groups between 5 to 54 years old, and a decrease in those groups aged older than 54 years old (Table 1).

### Prevalence

The crude prevalence of type 2 diabetes in the UK showed a marked increase from 2004 to 2021, rising from 2.95% to 5.41% of the population (Figure 2b and Table 2). The increase was not linear and appeared to be levelling-off slightly in later years (Figure 2b).

**Table 2.** Prevalence rate per 100 people by year, sex, and age groups.

| Group | 2004 | 2005 | 2006 | 2007 | 2008 | 2009 | 2010 | 2011 | 2012 | 2013 | 2014 | 2015 | 2016 | 2017 | 2018 | 2019 | 2020 | 2021 |
| --- | --- | --- | --- | --- | --- | --- | --- | --- | --- | --- | --- | --- | --- | --- | --- | --- | --- | --- |
| All UK | 2.95 | 3.17 | 3.37 | 3.56 | 3.73 | 3.9 | 4.07 | 4.22 | 4.35 | 4.55 | 4.73 | 4.86 | 5.00 | 5.09 | 5.18 | 5.27 | 5.31 | 5.41 |
| Male | 3.19 | 3.42 | 3.65 | 3.86 | 4.06 | 4.27 | 4.47 | 4.67 | 4.83 | 5.06 | 5.26 | 5.40 | 5.56 | 5.66 | 5.77 | 5.87 | 5.91 | 6.00 |
| Female | 2.71 | 2.91 | 3.10 | 3.26 | 3.39 | 3.53 | 3.66 | 3.78 | 3.88 | 4.04 | 4.20 | 4.31 | 4.45 | 4.51 | 4.59 | 4.67 | 4.71 | 4.82 |
| 0-4 | 0.00 | 0.00 | 0.00 | 0.00 | 0.00 | 0.00 | 0.00 | 0.00 | 0.00 | 0.00 | 0.00 | 0.00 | 0.00 | 0.00 | 0.00 | 0.00 | 0.00 | 0.00 |
| 5-15 | 0.01 | 0.01 | 0.01 | 0.01 | 0.01 | 0.01 | 0.01 | 0.01 | 0.01 | 0.01 | 0.01 | 0.01 | 0.01 | 0.01 | 0.01 | 0.01 | 0.01 | 0.01 |
| 15-24 | 0.05 | 0.06 | 0.06 | 0.07 | 0.07 | 0.08 | 0.08 | 0.08 | 0.09 | 0.09 | 0.10 | 0.10 | 0.11 | 0.12 | 0.12 | 0.12 | 0.12 | 0.13 |
| 25-34 | 0.25 | 0.26 | 0.28 | 0.3 | 0.31 | 0.33 | 0.35 | 0.36 | 0.38 | 0.40 | 0.43 | 0.44 | 0.46 | 0.48 | 0.49 | 0.51 | 0.52 | 0.56 |
| 35-44 | 0.95 | 1.02 | 1.10 | 1.17 | 1.24 | 1.33 | 1.41 | 1.49 | 1.56 | 1.65 | 1.72 | 1.76 | 1.83 | 1.86 | 1.90 | 1.93 | 1.95 | 2.00 |
| 45-54 | 2.87 | 3.09 | 3.29 | 3.46 | 3.62 | 3.77 | 3.92 | 4.05 | 4.18 | 4.40 | 4.58 | 4.70 | 4.89 | 5.01 | 5.16 | 5.28 | 5.36 | 5.53 |
| 55-64 | 5.83 | 6.19 | 6.56 | 6.92 | 7.27 | 7.67 | 8.08 | 8.42 | 8.70 | 9.09 | 9.40 | 9.62 | 9.89 | 10.03 | 10.14 | 10.24 | 10.32 | 10.47 |
| 65-74 | 10.46 | 11.16 | 11.8 | 12.28 | 12.60 | 12.91 | 13.22 | 13.38 | 13.47 | 13.81 | 14.11 | 14.33 | 14.66 | 14.88 | 15.11 | 15.41 | 15.63 | 15.79 |
| 75-84 | 10.67 | 11.75 | 12.74 | 13.65 | 14.49 | 15.32 | 16.07 | 16.71 | 17.26 | 18.03 | 18.64 | 19.05 | 19.53 | 19.74 | 19.79 | 19.79 | 19.73 | 19.58 |
| 85+ | 7.44 | 8.16 | 8.85 | 9.45 | 10.01 | 10.72 | 11.31 | 11.93 | 12.52 | 13.79 | 14.92 | 15.77 | 16.66 | 17.44 | 18.05 | 18.67 | 19.14 | 19.53 |

Again, there was evidence of a differential prevalence rate between the four nations of the UK. Northern Ireland consistently had the lowest prevalence rate – 2.3% in 2004, increasing to 4.96% in 2021 – and Wales had a notably higher prevalence rate, increasing from 3.71% in 2004 to 6.37% in 2021 (Figure 2b). The crude prevalence rate in England increased from 2.95% to 5.39% over the period. The pattern of the rate of increase in crude prevalence between the four nations of the UK was remarkably similar (Figure 2b).

The crude prevalence was higher in men than in women. In 2021 it was 6.00% in men versus 4.82% in women (Table 2). The crude, prevalence rate ratio of men to women increased from 1.18 in 2004 to 1.24 in 2021.

With the exception of the very elderly, 85+ years, the prevalence rate increased with increasing age. The prevalence rate increased over the observed time-period in all age groups. In 2004 the age group with highest age-specific prevalence rate was the 75 to 84-year-old group (Table 2), with a prevalence rate of 10.67%, and this increased to 19.58% in 2021. Always of interest is the number of younger people who are diagnosed with type 2 diabetes. The prevalence rate in the 35 to 44-year-old age group increased from 0.95% to 2.00% from 2004 to 2021, and from 0.25% to 0.56%, respectively, in those aged 25 to 34.

## Discussion

Although we used a novel, automated analytical platform to rapidly produce these metrics that would otherwise take months to produce using long-hand methods, this epidemiological study elicited some surprising findings. That the UK population prevalence of type 2 diabetes continued its seemingly inexorable increase was not a surprise. However, that the crude incidence of type 2 diabetes had not increased over almost 20 years was unexpected, but it transpired that this had been reported previously^7^. However, there was a shift to earlier onset and/or earlier diagnosis of type 2 diabetes. This was far more notable in women, with a mean, earlier diagnosis of five years. The mean age at diagnosis in men decreased by two years. It was evident that case ascertainment of type 2 diabetes during COVID19 in 2020 did have an impact, but incidence rates rebounded in 2021 back to essentially where it would have been expected to be and possibly higher due to delayed diagnosis during 2020. The temporary delay in diagnosis had little impact on the prevalence rates.

Interestingly, we report notable differences in incidence and prevalence of type 2 diabetes between UK nations, Northern Ireland producing the most favorable patterns, whilst Wales showed regrettably high rates of type 2 diabetes. Whilst incidence rates are always going to be more variable than prevalence rates, there was a period around 2004 to 2006 when the incidence rates varied markedly. This was probably due to the impact of the introduction of the Quality Outcomes Framework (QOF) in UK general practice. The introduction of QOF incentivized improved recording of diabetes and relevant clinical details in general practice. Overall disease burden remained higher in men.

The incidence of type 2 diabetes in the UK has been the subject of several studies, each shedding light on different aspects and time periods of the condition. One of our own studies reported the incidence of type 2 diabetes in the UK from 1991 to 2010 and revealed a significant increase in diagnosed cases. Specifically, the crude incidence rate in 2010 was 5.2 per 1,000 population [3.71/1,000] (corresponding findings from this study reported here in square parentheses). The study noted that the standardized incidence ratio increased across successive 5-year periods, indicating a rising trend. Additionally, the proportion of patients diagnosed under the age of 40 years increased over time, suggesting an earlier onset or earlier diagnosis of the disease^5^. Another study focused on the period from 2004 to 2014^8^. It found that while the prevalence rates of type 2 diabetes increased from 3.2% [3.0%] in 2004 to 5.3% [4.7%] in 2014, the incidence rates remained relatively stable throughout this period. This study also highlighted higher incidence and prevalence rates among men and in individuals from deprived socio-economic backgrounds. A more recent study covering 2009 to 2018 reported a decline in the incidence of clinically diagnosed type 2 diabetes. The incidence rate in men was reported to have decreased from 5.06 [4.48] per 1,000 person-years in 2013 to 3.6 [4.2] by 2018. Similarly, in women, the incidence rate fell from 4.5 [3.6] to 2.9 [3.4] per 1,000 person years over the same period. These studies collectively illustrate the dynamic nature of type 2 diabetes incidence in the UK, with variations in trends over different time periods, with an increase in younger age groups being diagnosed. Tate and colleagues also reported that the choice of clinical codes and data quality can affect the trends in the incidence of diabetes^7^.

The prevalence of type 2 diabetes in the UK has also been the subject of several studies, again revealing concerning trends. In a study referred to above, the crude prevalence rates of type 2 diabetes increased from 3.2% [3.0%] in 2004 to 5.3% [4.7%] in 2014^8^. Sharma and colleagues reported an increase in type 2 diabetes prevalence in the UK from 2.4% in 2000 to 5.3% [4.6%] in 2013^10^. We ourselves reported a crude prevalence rate of 5.4% [4.6%] in 2013^9^. In 2020, Diabetes UK reported a crude population prevalence of all diabetes in the UK of 7%, and that this rate was said to be broadly similar across the four constituent nations of the UK. It is unclear how they derived this value but it is believed that it came from a report from Public Health England^11^.

The relationship between incidence and prevalence can be complicated. Incidence can be stable whilst prevalence increases because of, say, a change in life expectancy following diagnosis because of improved care or treatment for example. Due to advances in the treatment of T2DM, and through improvements in the care of its related complications through statins and anti-hypertensives, improvements in life-expectancy for patients with T2DM may be disproportionately higher than for the population as a whole^12^. This may be evidence from this study which reports an increase in prevalence of T2DM in those aged 85+ from 7.44 to 19.53, whilst the incidence rate has actually decreased (7.92 to 6.39). The impact of hyperglycemia with regards to survival remains contentious^13^.

A plausible explanation for these differential findings between incidence and prevalence over 20 years is that the pool of people likely to ever be diagnosed with type 2 diabetes may be largely fixed, thus, the overall crude incidence remained the same but there were changes in age-specific incidence. The increasing prevalence may be due, therefore, to the increase in the typical duration of diabetes as a consequence of earlier onset and/or diagnosis, and possibly increased life expectancy in latter years. The clinical consequence in the community of earlier onset of type 2 diabetes, and increased diabetes duration, is likely to be an associated increase in diabetes-related complications. The earlier diagnosis in women is interesting. We checked to assure ourselves that it wasn’t related to gestational diabetes. We have no reason to believe that this is artefactual. This observation requires further thought and investigation but it would seem that much of the increased burden is related to this factor, although men too had two years earlier onset.

### Study limitations

The methods that we have used to identify cases of type 2 diabetes in previous studies were more sophisticated. For example, we also used exposure to medications specific to type 2 diabetes, and we had complicated algorithms to deal with the small number of cases who had diagnoses of both type 1 and type 2 diabetes and so on. Using the more detailed methods we used in previous studies the lifetime prevalence rates reported here – which have been validated^6^ – are likely to be essentially the same. Importantly, these methods are hard-coded and standardized in Livingstone, so that these analyses can be replicated repeatedly using identical methods, or with audited changes to the inputs.

Here we did not use linked data from hospital sources in this study because it would have precluded the generation of data from nations other than England. Again, knowing the nature of type 2 diabetes, we don’t expect this to have impacted our findings to any great extent. Inclusion of linked hospital data did not change the findings very much in our earlier epidemiological study^54^.

## Conclusions

The crude incidence of type 2 diabetes was stable over 20 years but there were changes over time in age-specific groups suggesting a shift to earlier onset and diagnosis in those who would eventually be diagnosed with the 2 diabetes. The prevalence of type 2 diabetes increased but the rate of increase appeared to be leveling-off just a little. There were notable differences in the epidemiology of type 2 diabetes between the four constituent countries of the UK. Most concerning was the high prevalence rate here in Wales. The mean age at diagnosis in women was reduced by more than five years, and around two years in men. A lot more younger people are being diagnosed with type 2 diabetes.

## Data Availability

CPRD data are available from the UK MHRA

https://cprd.com/

## Acknowledgements

* Livingstone Development Team: Christian A. Bannister, James Bateman, Thomas R. Berni, Aron Buxton, James J. Chesman, Craig J. Currie, Harry Fisher, Benjamin R. Heywood, Ellen Hubbuck, Sarah E. Holden, Bethan I. Jones, Sara Jenkins-Jones, Elgan R. Mathias, Christopher Ll. Morgan, Lauren D. Riddick, Christopher P. Shepherd, Darren R. Summers, Rhiannon K. Thomason, John Threlfall. There were no conflicts of interest.

---

**Supplementary Table1.**
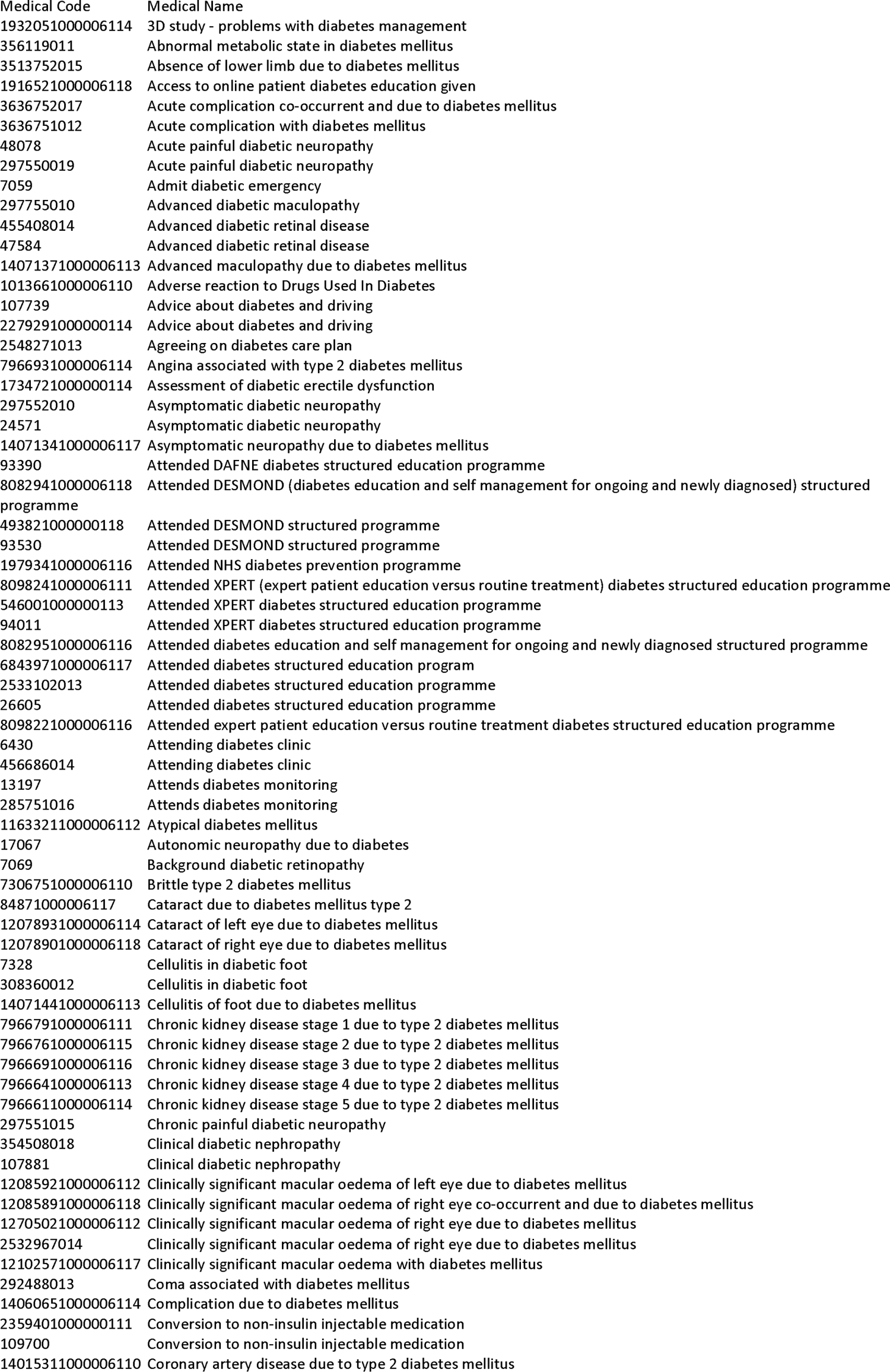
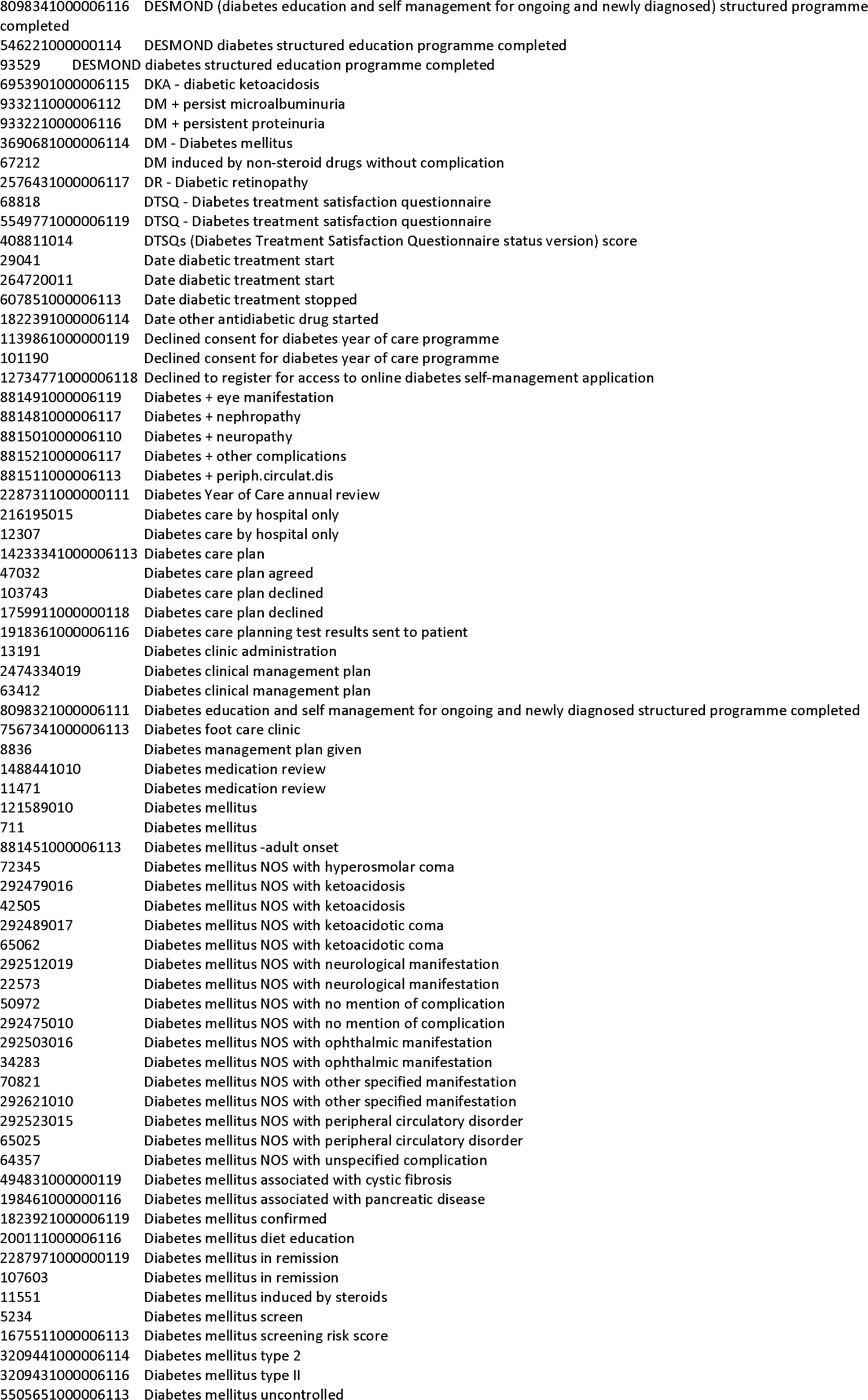
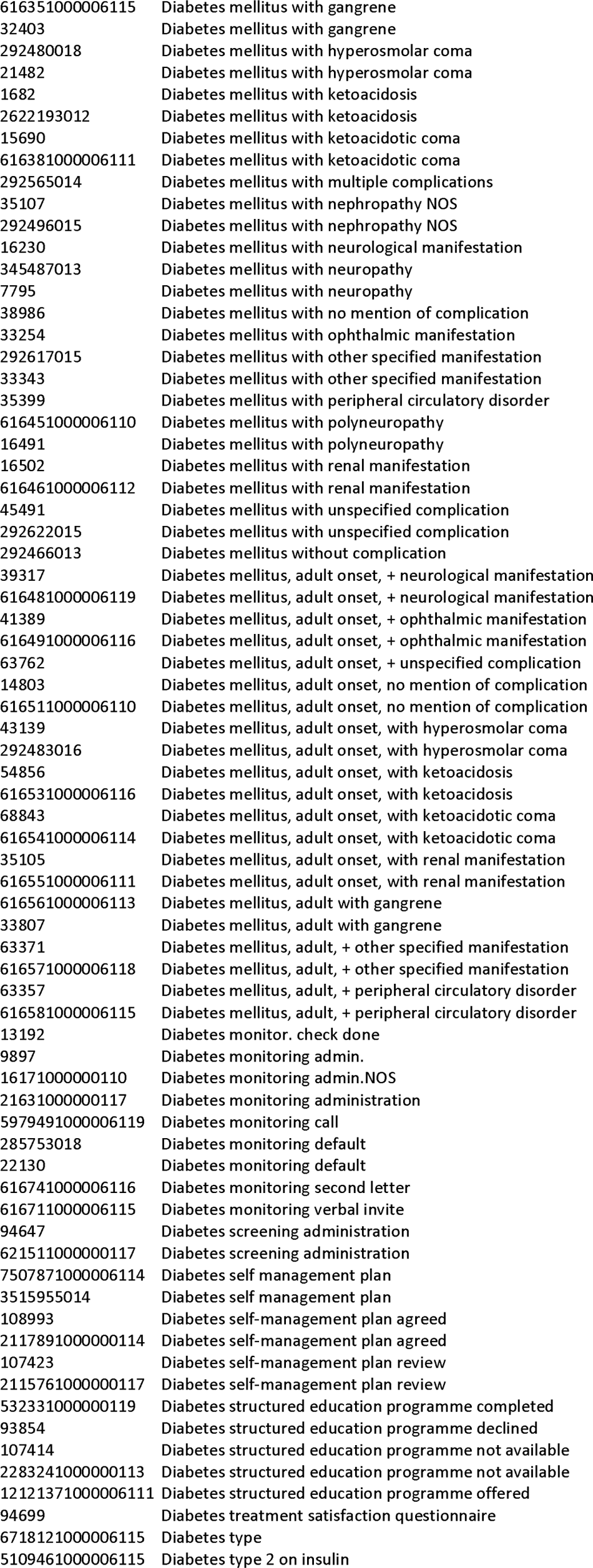
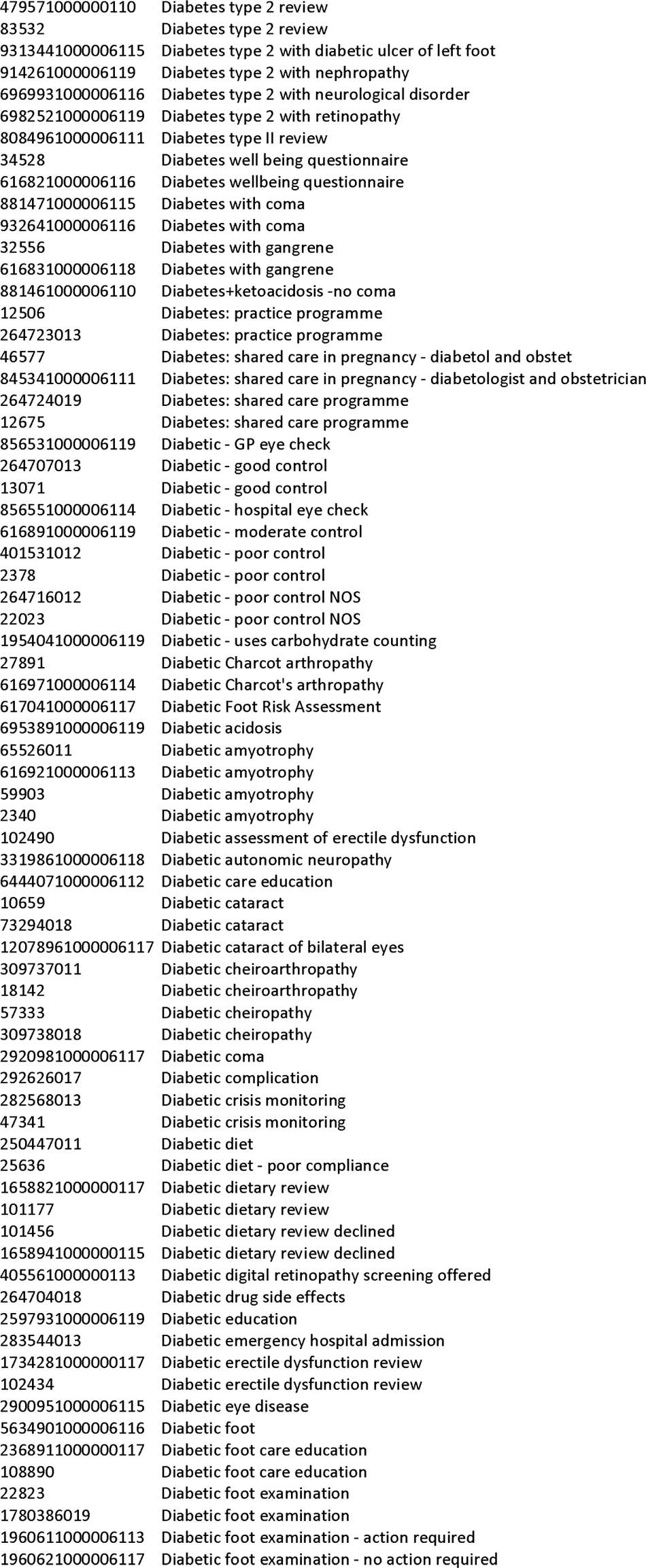
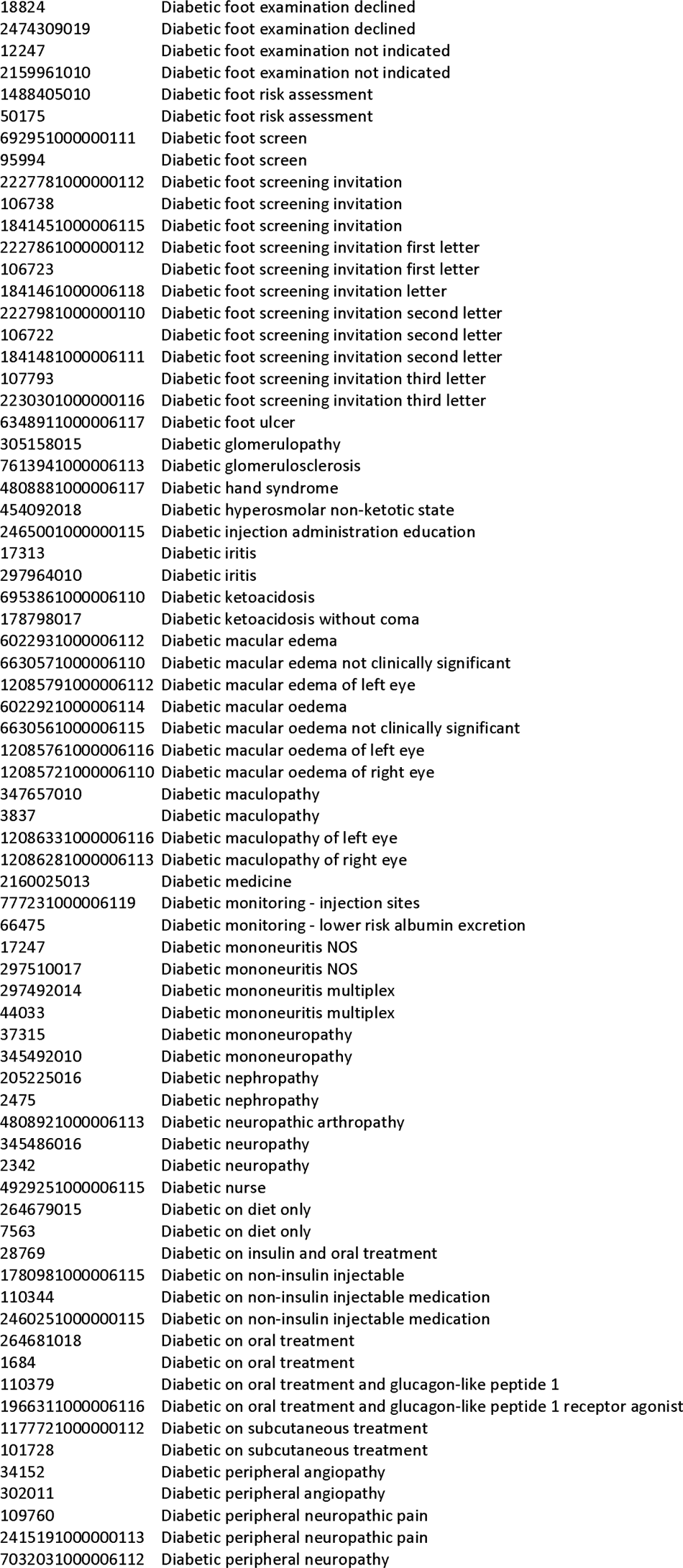
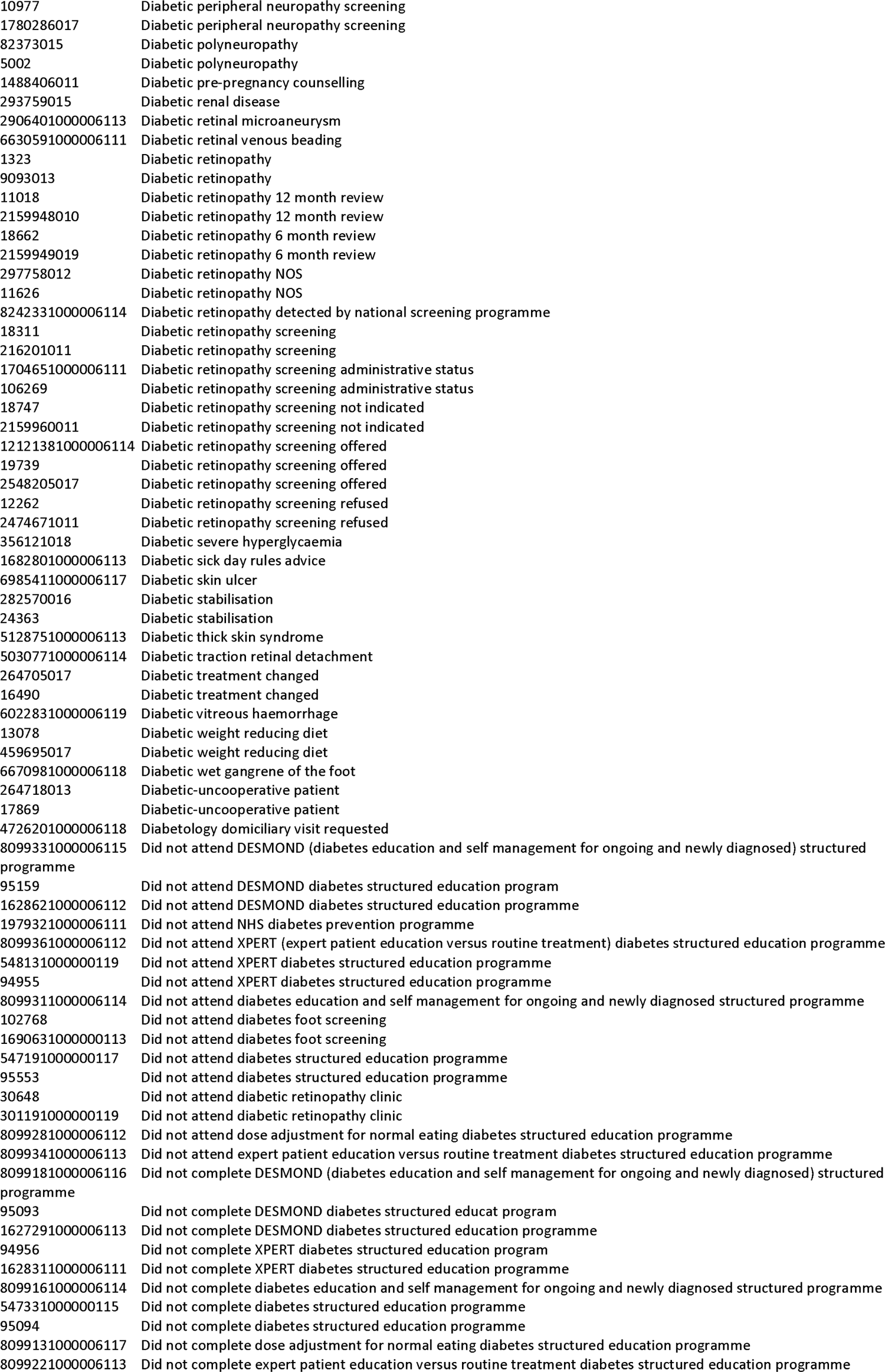
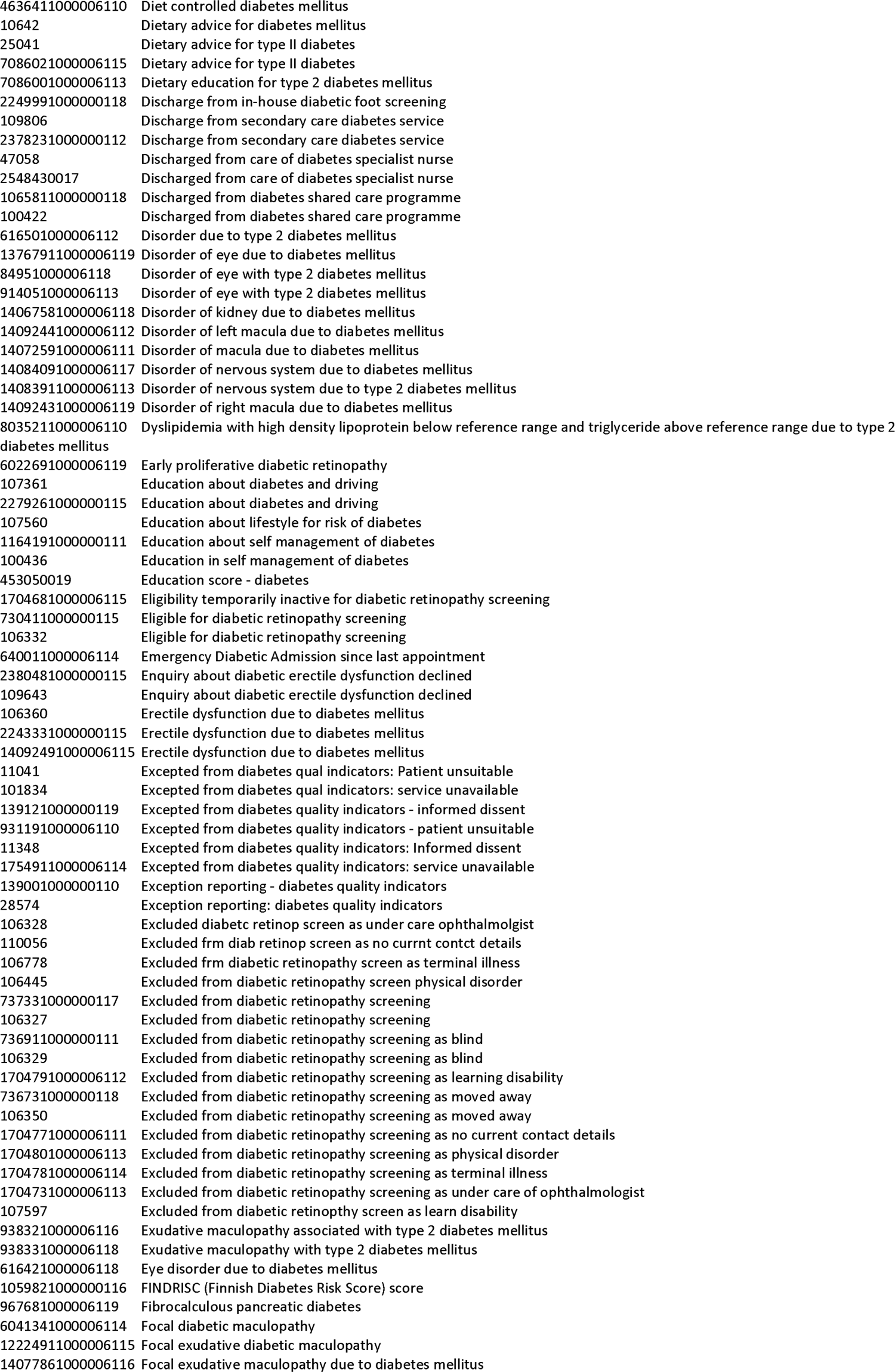
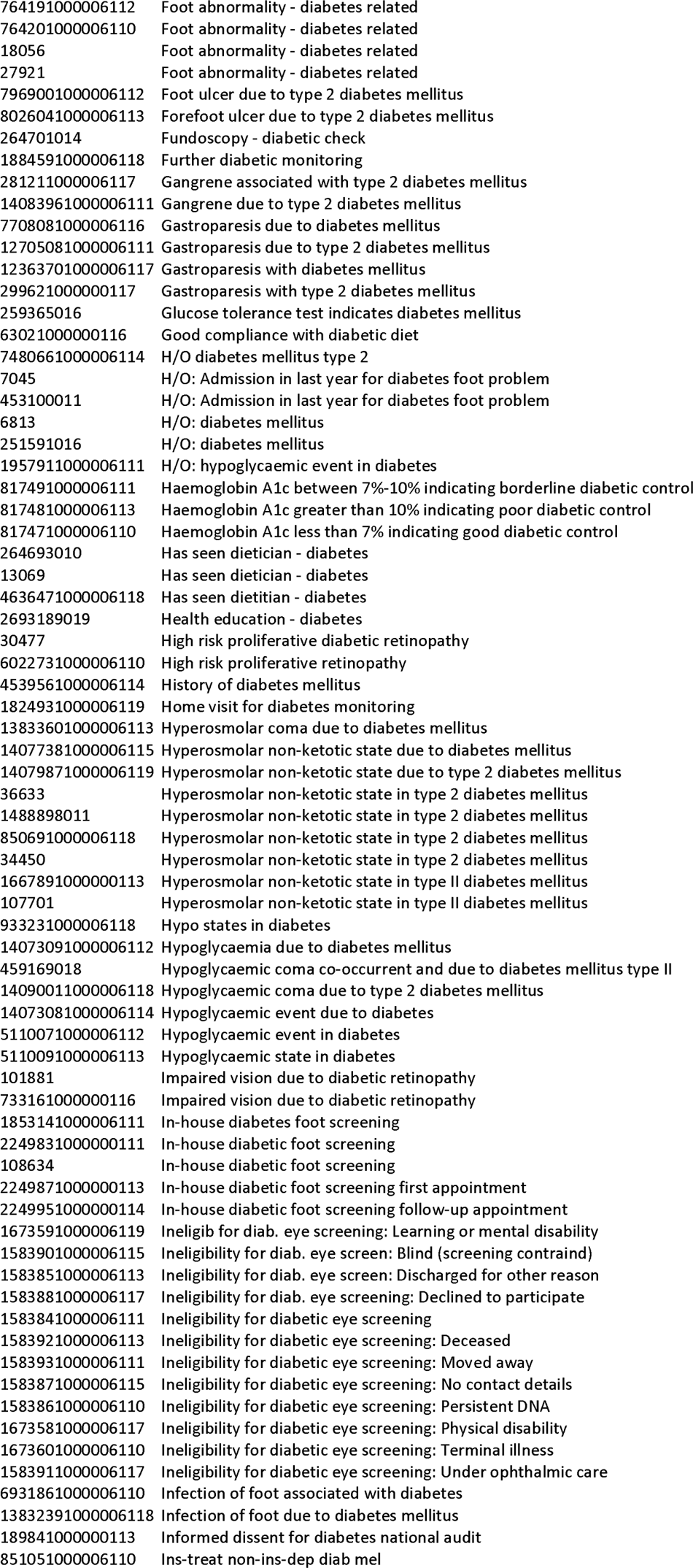
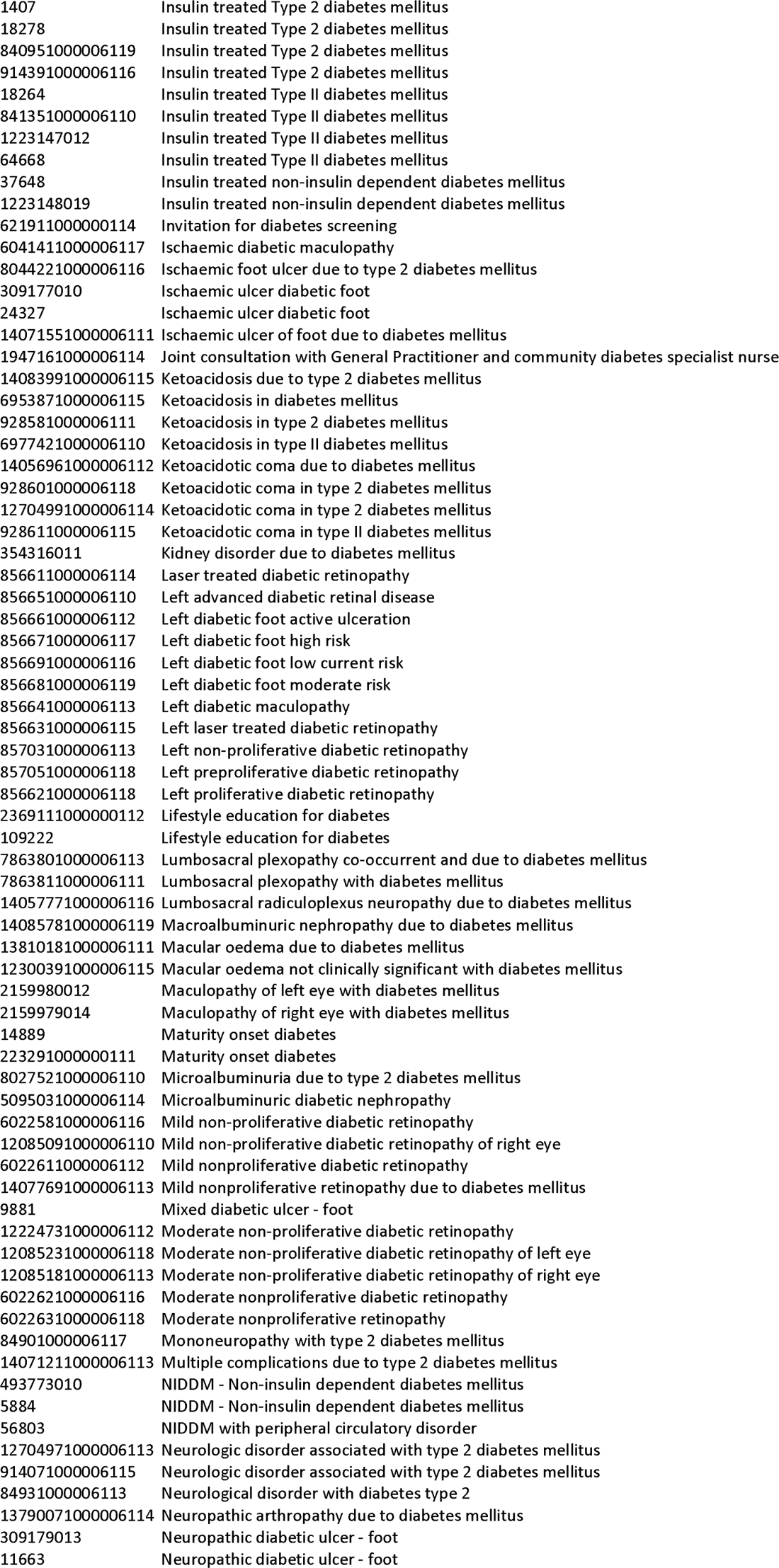
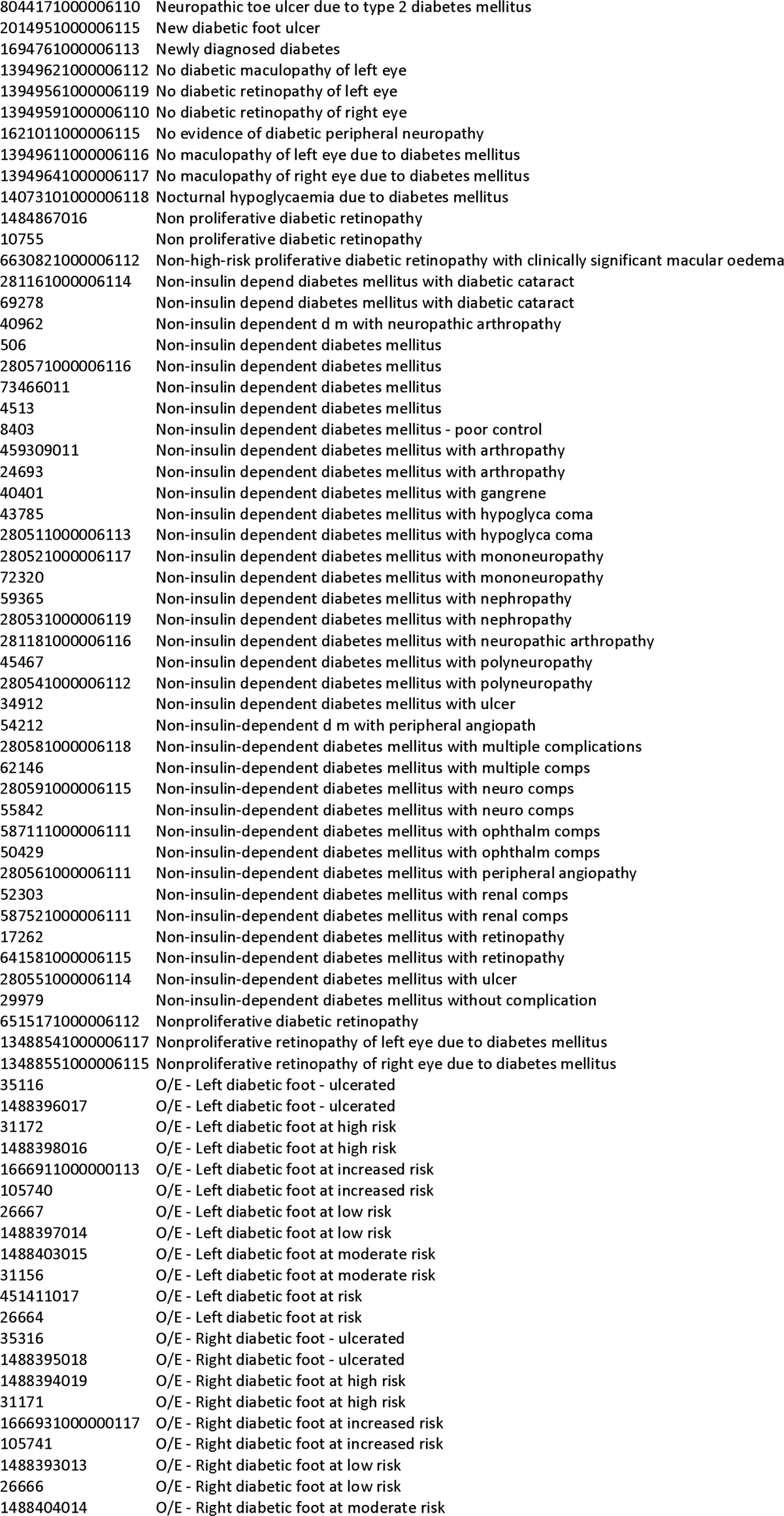
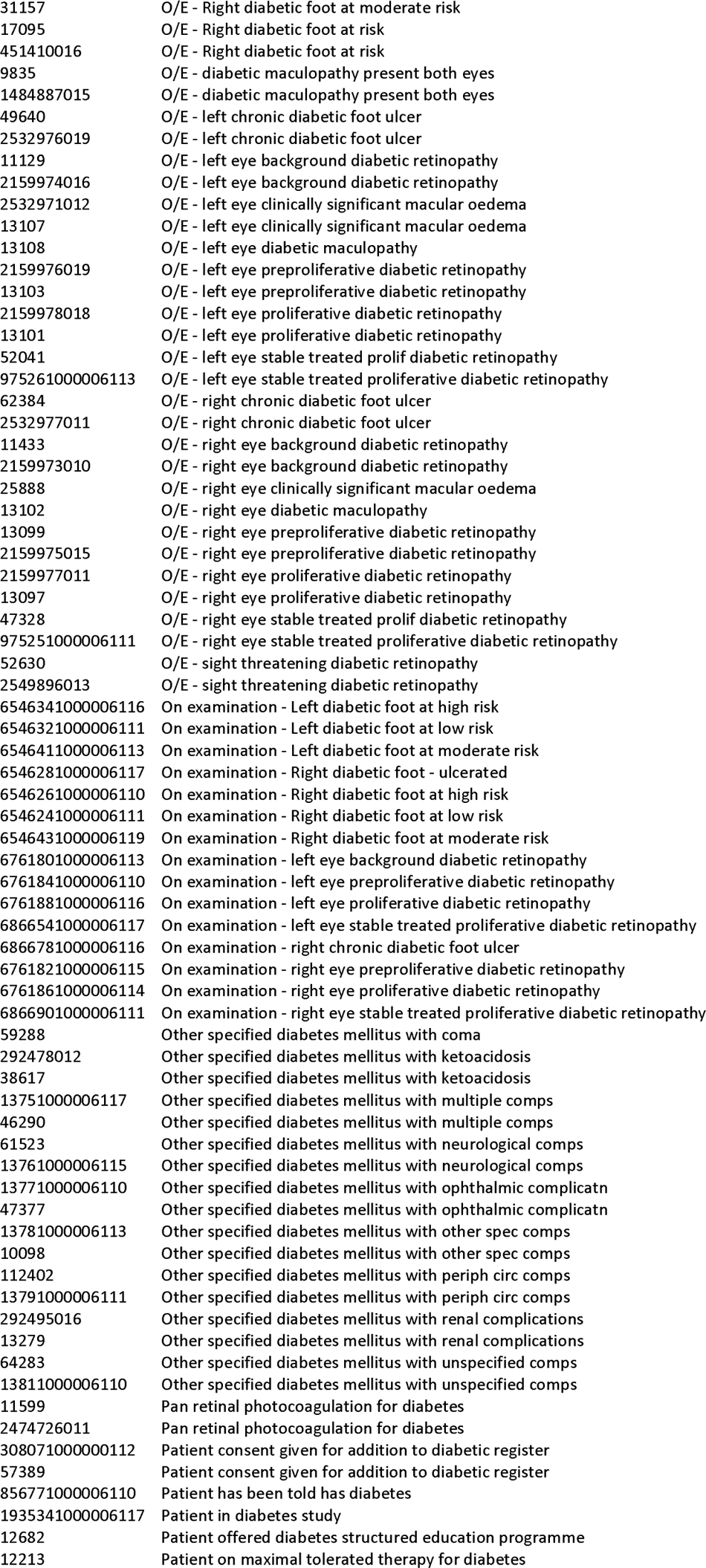
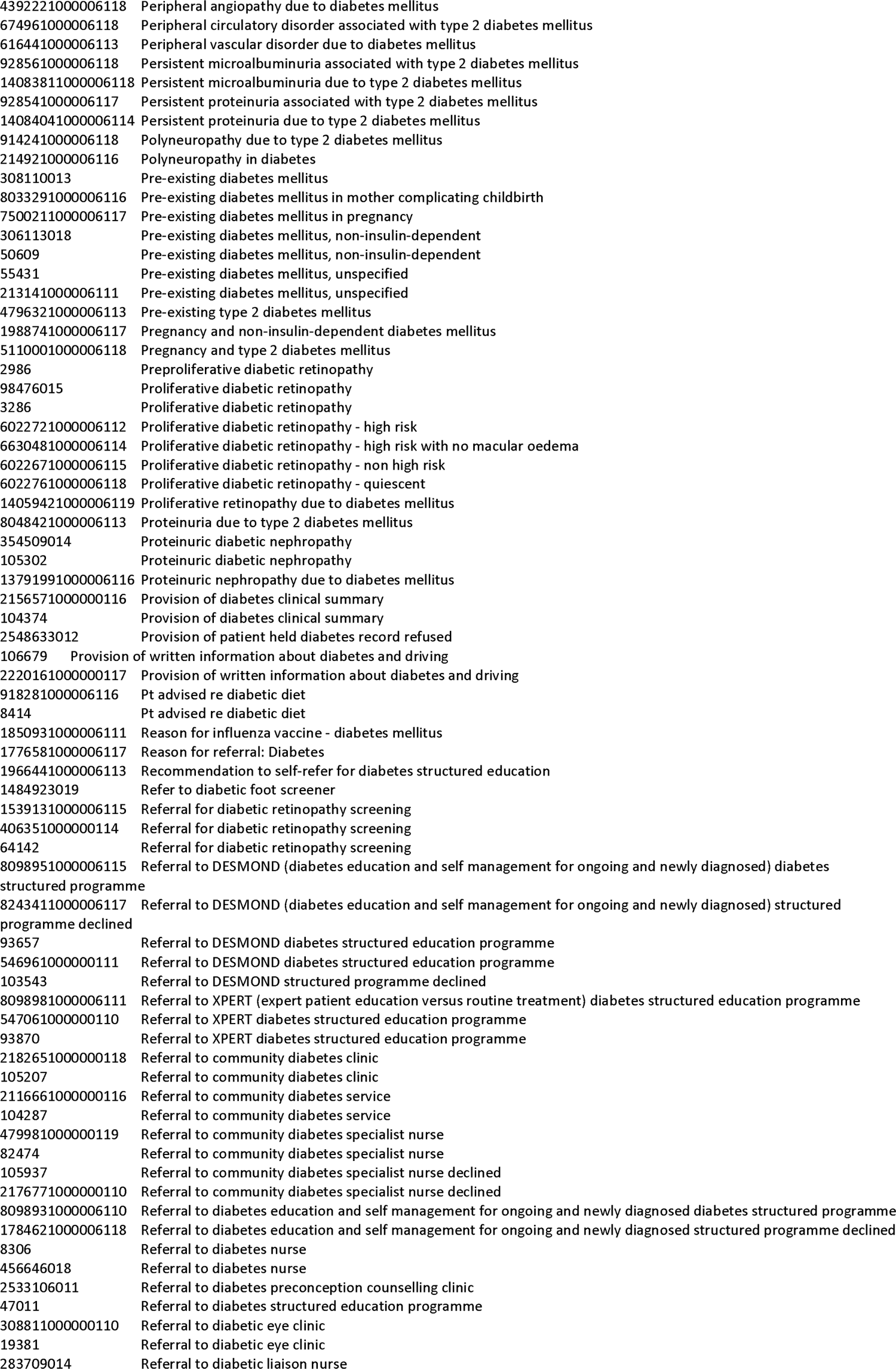
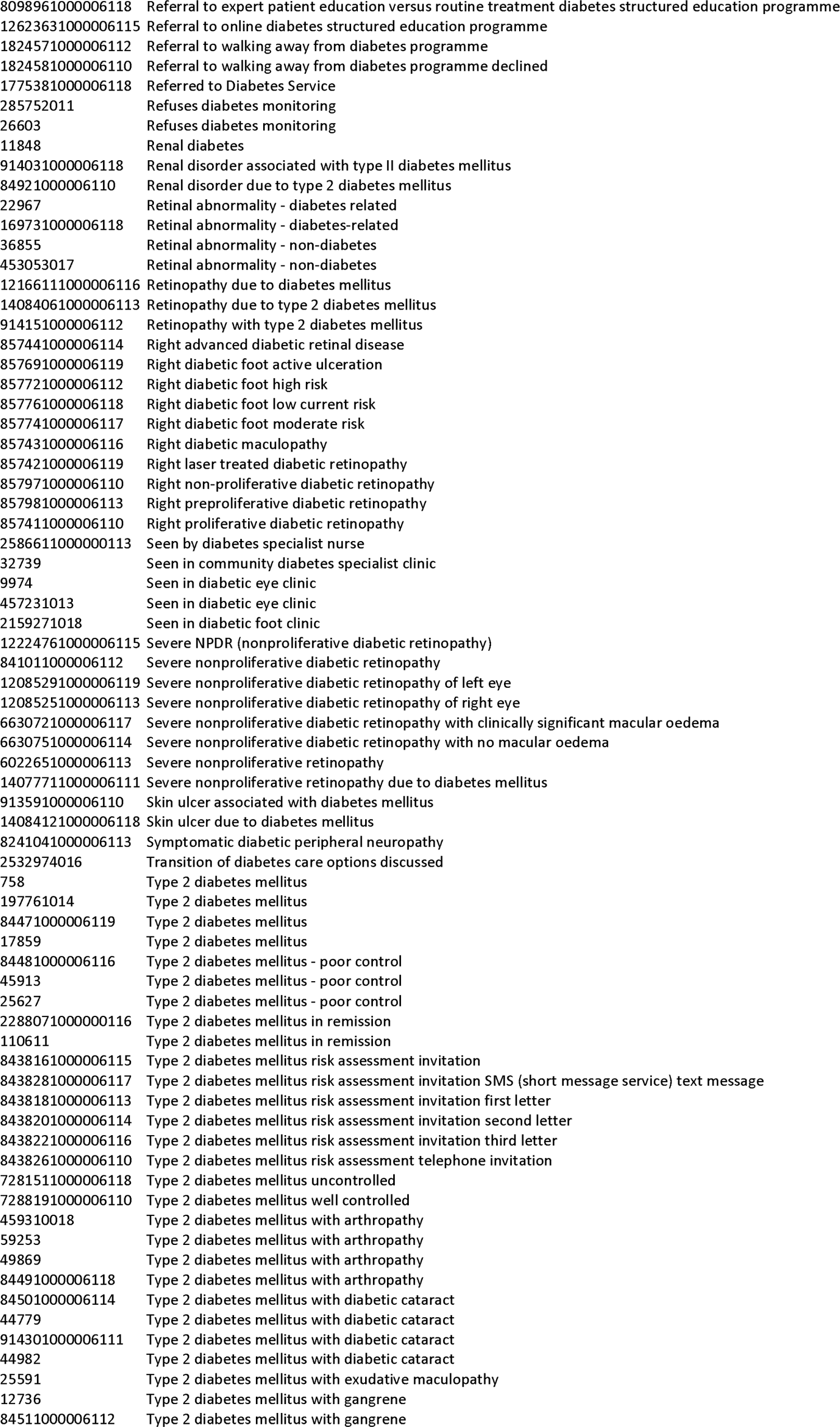
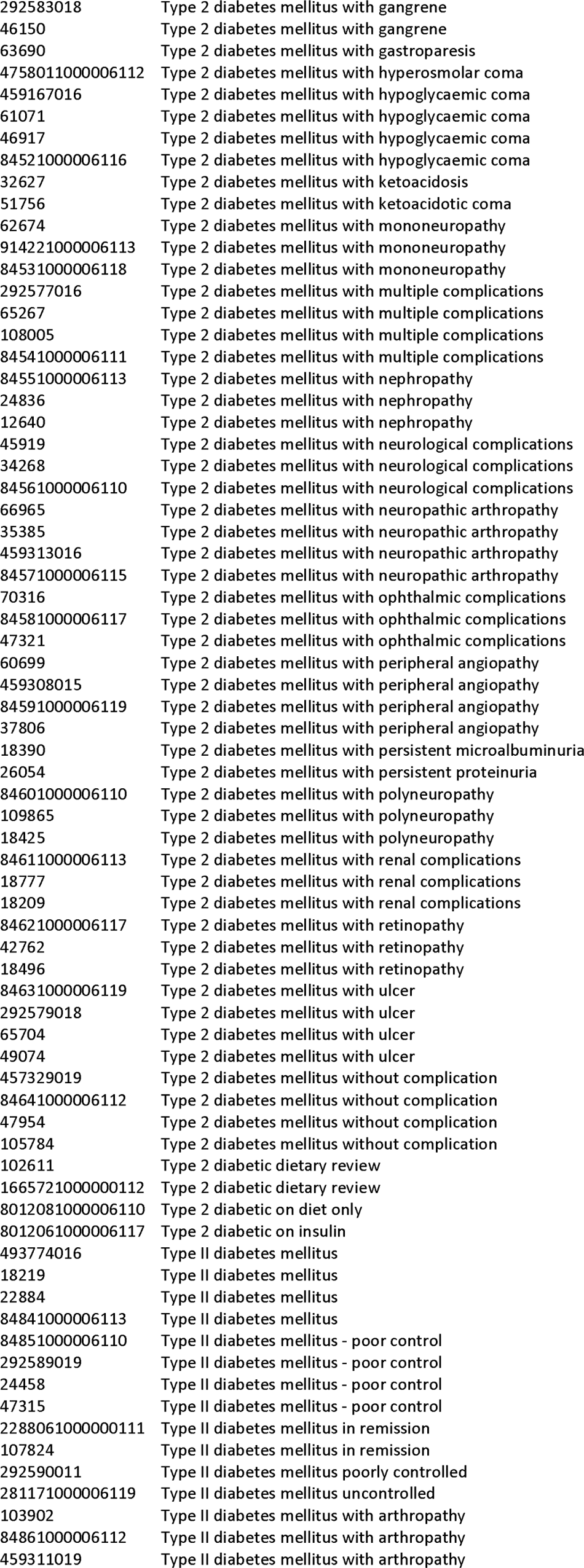
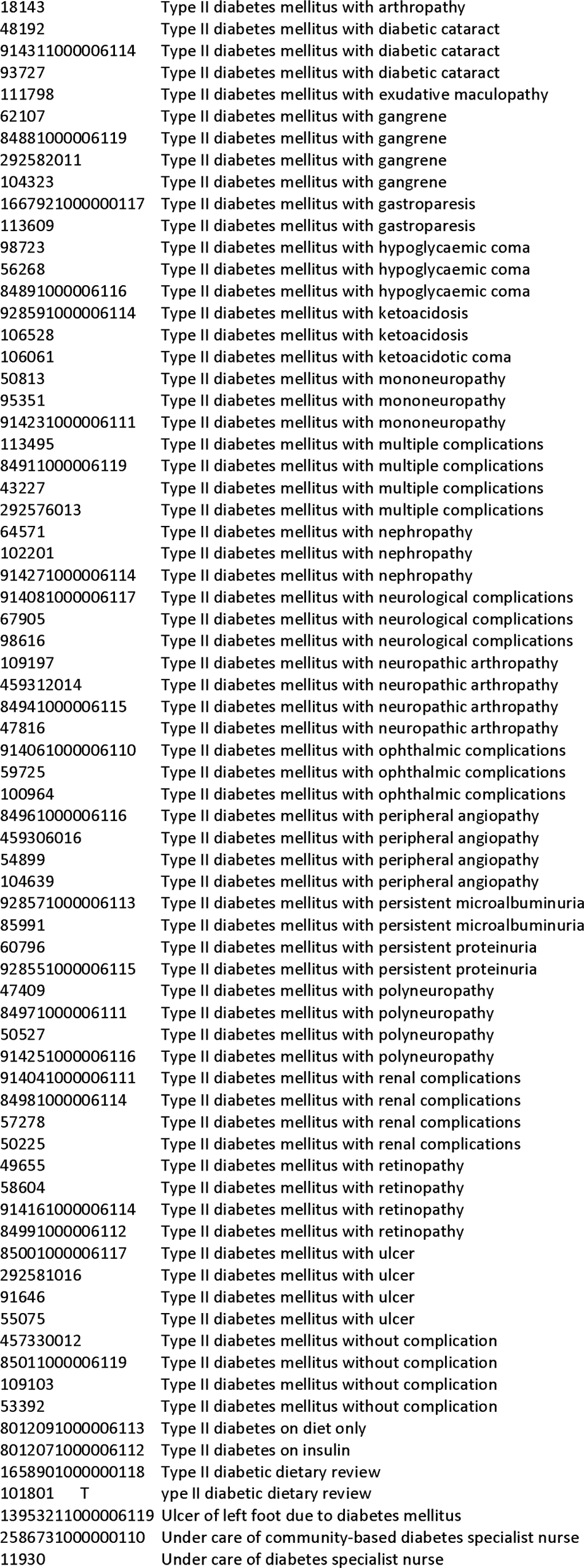
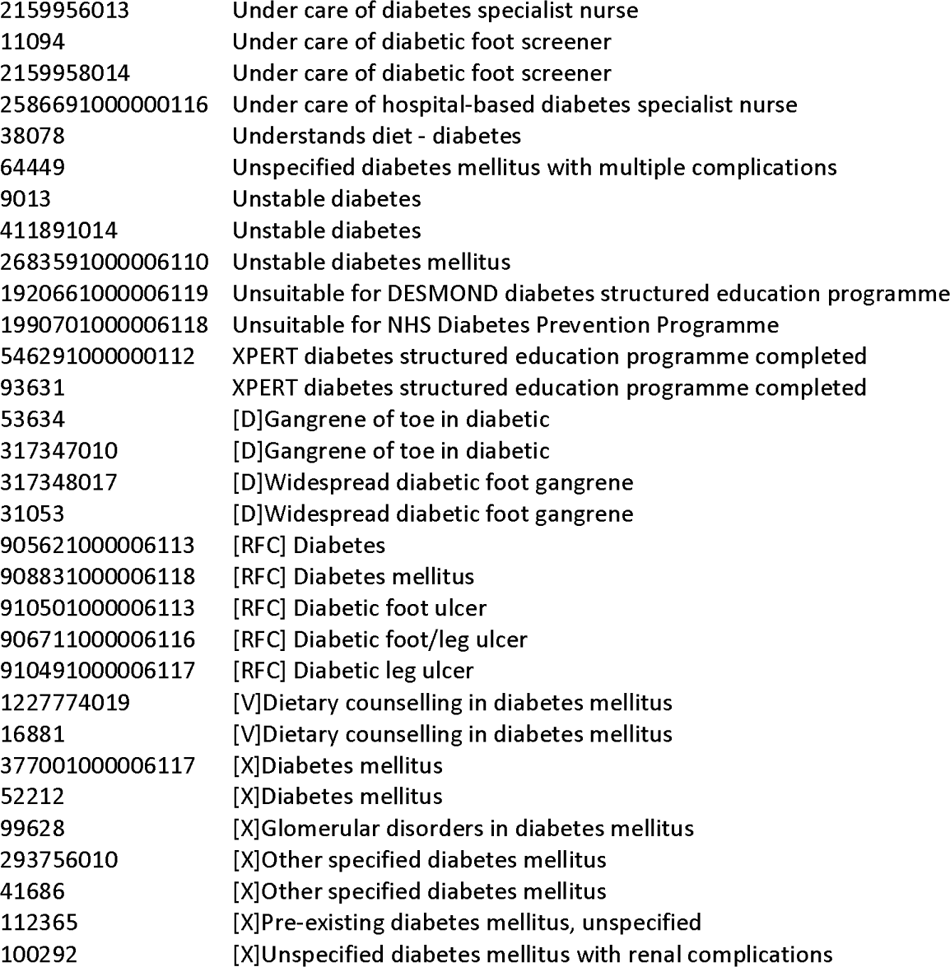
Medical codes used to identify type 2 diabetes.

## References

1. Evans JM, Barnett KN, Ogston SA, Morris AD. Increasing prevalence of type 2 diabetes in a Scottish population: effect of increasing incidence or decreasing mortality? Diabetologia. 2007;50:729–32

2. Office For National Statistics. National population projections: 2020-based interim. https://www.ons.gov.uk/peoplepopulationandcommunity/populationandmigration/populationprojections/bulletins/nationalpopulationprojections/2020basedinterim

3. Wang YC, McPherson K, Marsh T, Gortmaker SL, Brown M. Health and economic burden of the projected obesity trends in the USA and the UK. Lancet 2011;378:815–25

4. Holden SE, Jenkins-Jones S, Morgan CLl., Peters JR, Schernthaner G, Currie CJ. Prevalence, glucose control and relative survival of people with Type 2 diabetes in the UK from 1991 to 2013. Diabet Med 2017;34:770–780

5. Holden SH, Barnett AH, Peters JR, Jenkins-Jones S, Poole CD, Morgan CL, Currie CJ. The incidence of type 2 diabetes in the United Kingdom from 1991 to 2010. Diabetes Obes Metab 2013;15:844–52

6. Heywood BR, Morgan CLl., Berni TR, Summers DR, Jones BI, Jenkins-Jones S, Holden SE, Riddick LD, Fisher H, Bateman JD, Bannister CA, Threlfall J, Buxton A, Shepherd C, Mathias ER, Thomason RK, Hubbuck E, Currie CJ. Real-world evidence from the first online healthcare analytics platform-Livingstone. Validation of its descriptive epidemiology module. PLOS Digit Health 2023;2:e0000310

7. Tate AR, Dungey S, Glew S, Beloff N, Williams R, Williams T. Quality of recording of diabetes in the UK: how does the GP’s method of coding clinical data affect incidence estimates? Cross-sectional study using the CPRD database. BMJ Open 2017;25:e012905

8. Zghebi SS, Steinke DT, Carr MJ, Rutter MK, Emsley RA, Ashcroft DM. Examining trends in type 2 diabetes incidence, prevalence and mortality in the UK between 2004 and 2014. Diabetes Obes Metab 2017;19:1537–1545

9. Sharma M, Nazareth I, Petersen I. Trends in incidence, prevalence and prescribing in type 2 diabetes mellitus between 2000 and 2013 in primary care: a retrospective cohort study. BMJ Open 2016; 6:e010210

10. Whicher CA, O’Neill S, Holt RIG. Diabetes in the UK: 2019. Diabet Med 2020;37:242–247

11. Public Health England. Diabetes Prevalence Model. 2016. https://assets.publishing.service.gov.uk/media/5a82c07340f0b6230269c82d/Diabetesprevalencemodelbriefing.pdf

12. Gæde P, Oellgaard J, Carstensen B, Rossing P, Lund-Andersen H, Parving HH et al.Years of life gained by multifactorial intervention in patients with type 2 diabetes mellitus and microalbuminuria: 21 years follow-up on the Steno-2 randomised trial. Diabetologia. 2016;59:2298–2307

13. Currie CJ, Holden SE, Jones S, Morgan CLl., Voss B, Rajpathak SN, lemayehu B, Peters JR, Engel SS. Impact of differing glucose-lowering regimens on the pattern of association between glucose control and survival. Diabetes Obes Metab 2018;20:821–830

